# Diagnostic value of cutaneous manifestation of SARS-CoV-2 infection

**DOI:** 10.1101/2020.07.10.20150656

**Authors:** Alessia Visconti, Veronique Bataille, Niccolò Rossi, Justine Kluk, Ruth Murphy, Susana Puig, Rabi Nambi, Ruth C. E. Bowyer, Benjamin Murray, Abigail Bournot, Jonathan Wolf, Sebastien Ourselin, Claire J. Steves, Tim D Spector, Mario Falchi

## Abstract

**Importance:** SARS-CoV-2 causes multiple immune-related reactions at various stages of the disease, and the wide variety of cutaneous presentations has delayed linking these to the virus. Previous studies had attempted to look at the prevalence and timing of COVID-19 rashes but were mostly based on hospitalized severe cases with limited follow up.

**Objective:** To assess the diagnostic value of new skin rashes in SARS-CoV-2 infection.

**Design:** Observational study including data collected longitudinally *via* the COVID Symptom Study app between May 7^th^ and June 22^nd^, 2020, as well as data from an independent online survey on skin-related symptoms.

**Setting:** Community-based

**Participants:** Volunteer sample of 336,847 UK users of the COVID Symptom Study app and 11,546 surveyees, aged 1 to 90 years old.

**Exposure:** Users self-reporting a positive or negative SARS-CoV-2 swab test result, untested symptomatic users, and survey respondents.

**Main Outcome(s) and Measure(s):** the diagnostic value of new skin rashes in SARS-CoV-2 infection, and observation on their duration and timing in relation to other COVID-19 symptoms

**Results:** In the app data, 8.8% of the swab positive cases (N=2,021) reported either a body rash or an acral rash, compared to 5.4% of those with a negative swab test (N=25,136). Together, these two cutaneous presentations showed an odds ratio (OR) of 1.67 (95% confidence interval [CI]: 1.42-1.97) for being swab positive. Skin rashes were also predictive in the larger untested group of symptomatic app users (N=54,652), as 8.2% of those who had reported at least one classic COVID-19 symptom, *i*.*e*., fever, persistent cough, and/or anosmia, also reported a rash. Data from the independent online survey showed that in 17% of swab positive cases, the rash was the initial presentation. Furthermore, in 21%, the rash was the only clinical sign.

**Conclusions and Relevance:** Skin rashes cluster with other COVID-19 symptoms, are predictive of a positive swab test and occur in a significant number of cases, either alone or before other symptoms. Recognising rashes is important for the early detection of COVID-19 cases. To help healthcare professionals in this task we have established a large library of high-quality manually curated photos, available at: https://covidskinsigns.com

**Key points:** 

**Question:** What is the diagnostiv value of the cutaneous manifestation of SARS-CoV-2 infection?

**Findings:** We confirmed, in a community-based setting, that the presence of a rash is predictive of SARS-CoV-2 infection, and provided a large library of cutaneous manifestation photos to help healthcare professionals in diagnosing COVID-19.

**Meaning:** Skin rashes should be considered as part of the clinical presentation of COVID-19 to aid earlier diagnosis and curb the spread of the infection.

## Introduction

During the COVID-19 pandemic, it became clear that the SARS-CoV-2 virus, whilst mainly targeting the lungs, also affected other organs^1^. Cutaneous manifestations were slower to be reported, possibly because in patients in critical conditions the need for documenting cutaneous changes was less pressing, and because the diversity of the associated skin changes may have delayed their initial association with COVID-19 infection^2^. The first cases of COVID-19 cutaneous manifestation were documented in China, but prevalence was very low at 0.2% in 1,099 hospital cases^3^. Italy then reported that 20% of the patients on a COVID-19 ward (N=88) had cutaneous signs^4^. Subsequently, other groups^2,5–9^ have described urticarial rashes, vesicular lesions and less frequent cases of chilblains affecting fingers or toes (acral rash), thought to be due to minor thrombotic events or damage to the endothelial walls of small distal vessels of the digits. Here, using a population approach, we investigated the diagnostic value of body and acral rashes for SARS-CoV-2 infections using data from 336,847 users of the COVID Symptom Study app, and from an independent survey on COVID-19-related cutaneous symptoms in 11,546 subjects, 2,328 of whom also shared photos of their skin complaints.

## Methods

### The COVID Symptom Study app

The COVID Symptom Study app^10^ was developed by Zoe Global Limited, supported by physicians and scientists at King’s College London and Massachusetts General Hospital, Boston, and was first launched in the UK on March 24^th^, 2020. Participants were recruited through social media outreach or invitations from the investigators of long-running cohort studies to their volunteers within the newly established COronavirus Pandemic Epidemiology (COPE) Consortium^10^. The study population includes anyone able to download the app, either themselves or by proxy. The app collects on sign up, data on sex, age, ethnicity (*i*.*e*., Asian, Black, Chinese, Middle East, Mixed, or White), core health risk factors, including height, weight, and common disease status (*e*.*g*., cancer, diabetes, heart, kidney, and lung disease), the use of medications (*e*.*g*., corticosteroids, immunosuppressants and blood pressure medications). It also records, whether the user is a healthcare worker. From May 7th, 2020, users were prompted to self-reportif they had ever had a SARS-CoV-2 test. how it was performed (*e*.*g*., nose/throat swab, antibody testing), and the result (*i*.*e*., positive, negative, failed, or waiting). Users could provide daily updates on their health status and the presence of up to 14 COVID-19-related symptoms: abdominal pain, chest pain, delirium, diarrhoea, fatigue, fever, headache, hoarse voice, anosmia, persistent cough, shortness of breath, skipped meals, sore throat, and unusual muscle pains. From April 29th, 2020, two cutaneous manifestations were also added: raised, red, itchy wheals on the face or body or sudden swelling of the face or lips (body rash), and red/purple sores or blisters on the feet or toes (acral rash).

### Study population

This study included residents in the UK aged from 1 to 90 years who downloaded the app and entered regular data between May 7^th^ and June 22^nd^, 2020, either themselves or *via* proxy. We excluded users with body mass index (BMI) outside the range of 15 to 55 kg/m^2^ (for users 16 years old or older) or outside two standard deviation from the sample’s mean for each age (for users younger than 16 years old), pregnant women, and users who did not report their sex (**Supplementary Figure 1**). We removed inconsistent daily assessments, such as those with a logged body temperature outside the range of 35 to 43°C, or where users reported feeling unwell but had no symptoms. This resulted in 336,847 users, 17,407 of whom also provided valid (*i*.*e*., positive or negative) results for SARS-CoV-2 swab tests (hereafter: “tested users”). We further selected 54,652 symptomatic users (*i*.*e*., users reporting at least one of the 16 collected symptoms during their daily log history) who did not consider themselves of having been already infected when first registering with the app and were not tested *via* nose/throat swab (hereafter: “symptomatic untested users”). These were divided in two groups: those reporting at least one of the of the three main symptoms of COVID-19 (*i*.*e*., fever, persistent cough, and/or anosmia, hereafter: “classic symptoms”) either at the time of logging or in retrospect, and who, according to the NHS guidelines, would require isolation and testing, and those who did not (**Supplementary Figure 2**).

**Figure 1.**
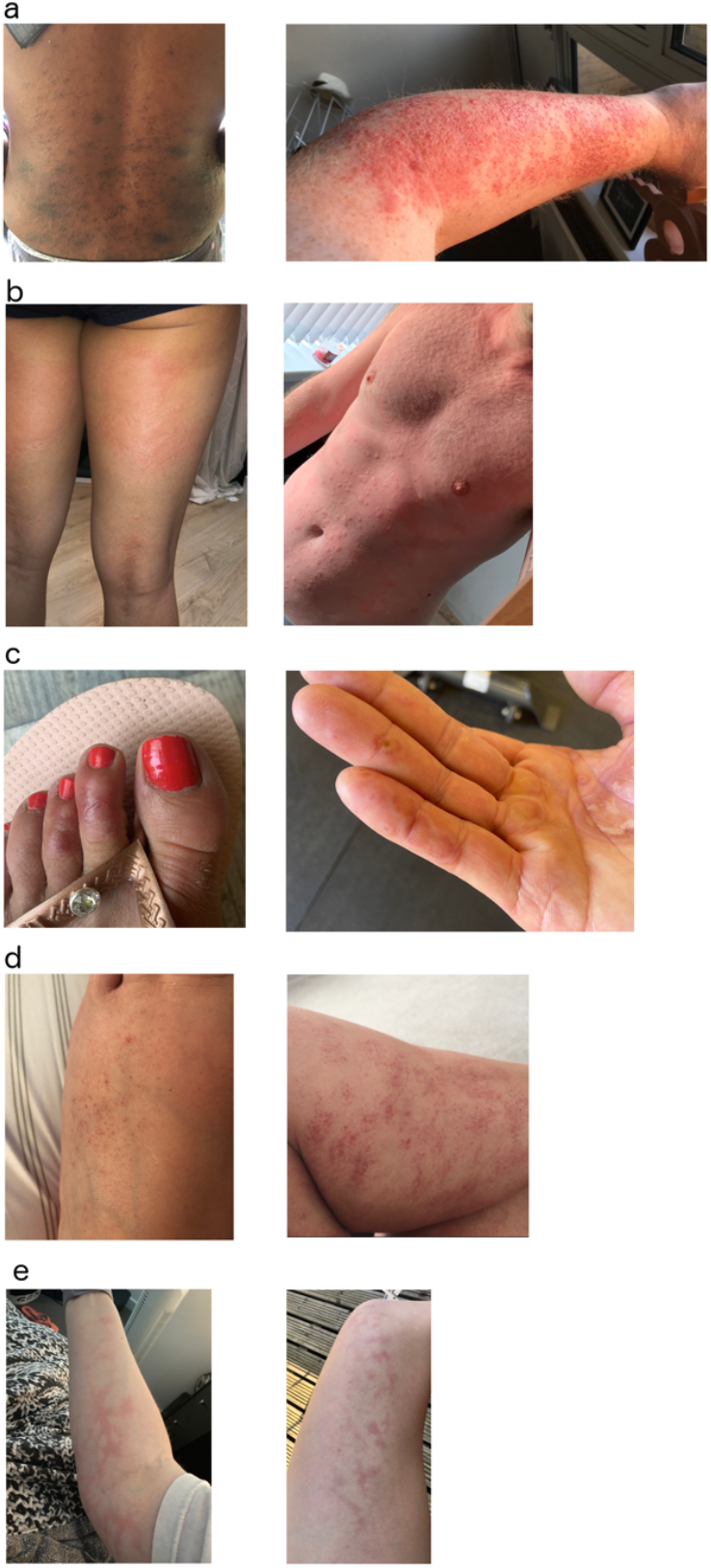
Example of COVID‐19 related cutaneous manifestations. **(a) Papular rash**. Right: erythematopapular rash on the back. Left: erythematopapular eruption on the forearm, some blistering and necrosis of the top layers of the epidermis is also visible. **(b) Urticarial rash**. Right: large urticated plaques on the back of thighs and popliteal fossae. Left: widespread urticaria on torso **(c) Acral rash**. Left: erythema on the dorsal aspect of the second and third toe with a blister on the second toe. Right:erythematous annular lesions with some shedding of the epidermis on fingers and palms **(d) Vasculitic body**. Right: petechiae on the dorsum of foot. Left: multiple petechiae with blood cells extravasation on calf. **(e) Livedo reticularis**. Right: livedo reticularis on arm. Left: livedo reticularis on thigh.

**Figure 2.**
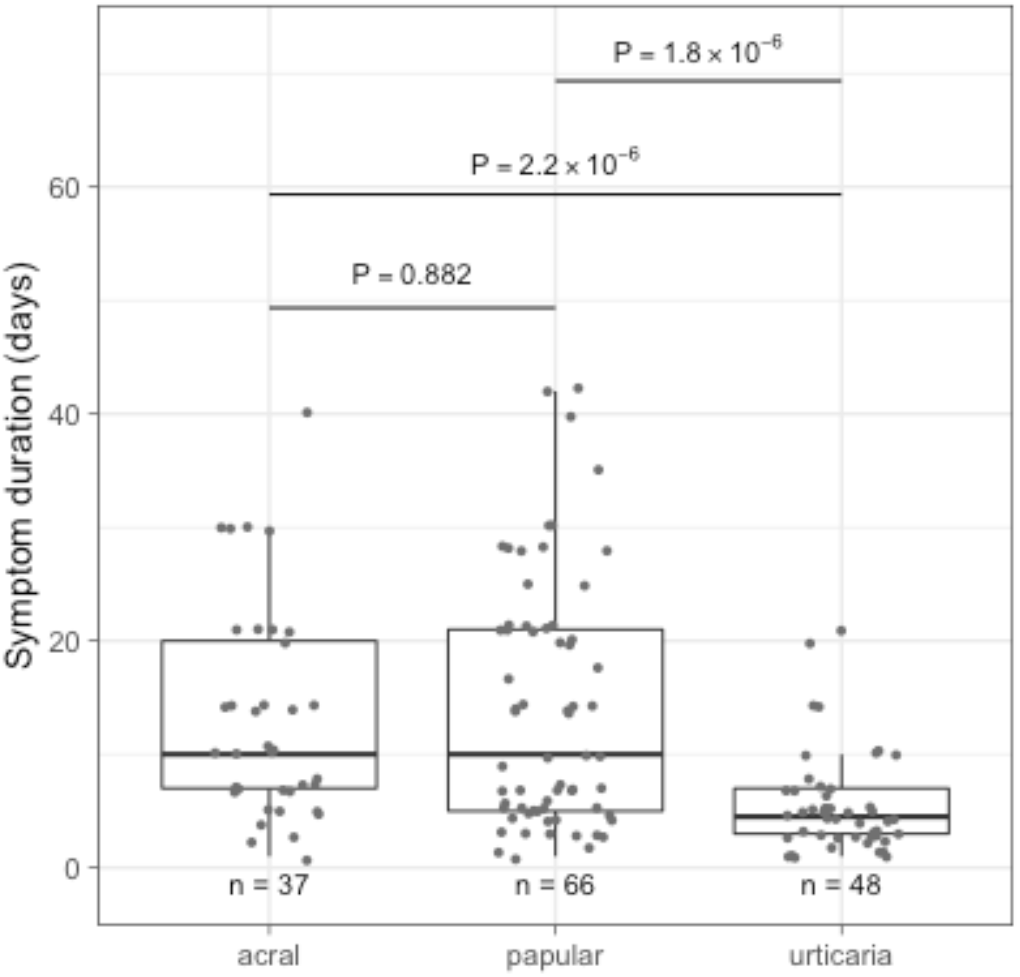
Duration of symptoms. Distribution of symptom duration is shown for the three most common cutaneous symptoms diagnosed from the selected photos. P values were generated by means of the Wilcoxon’s test.

### The skin rash survey

To collect more detailed information on body and acral rash duration and timing with respect to other COVID-19 symptoms, and to create a repository of photos for COVID-19-related cutaneous symptoms, we delivered an independent online questionnaire *via* Survey Monkey, asking whether the rash was the only symptom, how many days it lasted, and, if other COVID-19-related symptoms were present, whether the rash started before, during or after them. Participants were recruited through social media outreach.

The questionnaire was open from 12^th^ to 17^th^ June, 2020, and 29,966 individuals participated. We removed 18,422 surveyees not reporting their sex, age, the duration of their cutaneous symptoms, or reporting symptoms lasting more than six weeks, or being outside the 1-90 years old range (**Supplementary Figure 3)**. Of the 11,544 surveyees passing this quality control, 2,328 uploaded a photo of their rash, and gave consent for sharing. From these, we randomly selected 260 photos from both sexes, with either a positive SARS-CoV-2 test or reporting at least one of the three classic symptoms. The 260 photos were blindly assessed and independently categorised by four experienced dermatologists. Classifications were accepted when at least three dermatologists agreed on the diagnosis.

In collaboration with the British Association of Dermatologists, the complete photo database was subsequently examined by four consultant dermatologists, and about 400 photos were classified by consensus to create a large library of curated photos divided in several types of rashes.

### Ethical statement

The study has been approved by the King’s College London Research Ethics Committee REMAS ID 18210, review reference LRS-19/20-18210. All app users and surveyees provided informed consent, either themselves or by proxy. Additional consent was sought for the sharing of the uploaded photos with researchers, healthcare professionals, and journalists for publication purpose.

### Statistical analyses

Statistical analyses were carried out using R (v3.6.1). To identify confounders, comparisons between categorical and continuous values were carried out using Pearson’s χ^2^ test and Wilcoxon’s test, or linear regression, respectively. Associations between the presence/absence of self-reported skin-related symptoms and, in tested users, SARS-CoV-2 test results, and, in symptomatic untested users, the presence/absence of the three classic COVID-19 symptoms, were carried out through multivariate logistic regression, and the following variables, identified in the previous analysis, were included as covariates: sex, age, BMI, ethnicity, smoking status (never, ex, current), common disease status (diabetes and lung disease) and whether corticosteroids, immunosuppressants, or blood pressure medications were administered. Associations passing a Bonferroni-derived threshold of 0.05/2 (body rash and acral rash)=0.025 were considered as significant. Sensitivity analyses were carried out by stratifying for sex, ethnicity, and being a healthcare worker.

## Results

The majority of the 336,847 UK users of the app who registered between May 7th and June 22^nd^ 2020 and were included in this study were white European (94.0%). Results for SARS-CoV-2 swab tests were provided by 27,157 users (8.1%), 2,021 of whom (7.4%) were positive. Among untested users, 54,652 were symptomatic (*i*.*e*., reported at least one of the 16 collected symptoms), including 17,371 users presenting with at least one of the three symptoms of COVID-19 (*i*.*e*., fever, persistent cough, and/or anosmia) whose presence, according to the UK NHS guidelines, would require isolation and testing. While these guidelines are not diagnostic, the presence of any of these three symptoms associates in our data with SARS-CoV-2 positive swab with an odds ratio (OR) of 5.69 (95% Confidence Interval [CI] = 5.13-6.31, P=5.12×10^−282^), indicating with good confidence an enrichment for infected subjects. Sample characteristics are summarised in **Table 1** and **Supplementary Table 1**

**Table 1.** Sample characteristics. Categorical values are reported as number and percentage, and compared using Pearson’s χ^2^ test. Continuous values are reported as mean ± standard deviation and compared using Wilcoxon’s test. Associations P values with body and acral rash are from multivariate logistic regression, adjusted for the relevant covariates (**Methods**), and with BMI from multivariate linear regression, adjusted age and sex. “Users tested **for SARS-CoV-2**” refers to users self-reporting a positive or negative swab test result. “Symptomatic untested users” refers to users who reported at least one of the 16 collected symptoms, did not believe that they had already had COVID-19 when first registering with the app, had not yet been tested for SARS-CoV-2. “Classic symptoms” refers to those included in the NHS guidelines (*i*.*e*., fever, persistent cough, and/or anosmia).

|  | All users | Users tested for SARS-CoV-2 |  |  |  | Symptomatic untested users |  |  |  |
| --- | --- | --- | --- | --- | --- | --- | --- | --- | --- |
|  |  | All | Positive | Negative | P value | All | With classic symptoms | Without classic symptoms | P value |
| **N** | 336,847 | 27,157 | 2,021 | 25,136 | - | 54,652 | 17,371 | 37,281 | - |
| **Females (%)** | 188,118<br>(55.8%) | 16,474<br>(60.7%) | 1,376<br>(68.1%) | 15,098<br>(60.1%) | 1.47x10<sup>-12</sup> | 34,789<br>(63.7%) | 10,684<br>(61.5%) | 24,105<br>(64.7%) | 1.03x10<sup>-12</sup> |
| **Age** | 43.9±19.7 | 43.9±17.5 | 43.9±15.6 | 43.9±17.7 | 0.09 | 41.4±18.5 | 38.2±19.4 | 42.9±17.8 | 1.51x10<sup>-145</sup> |
| **BMI** | 26.2±6.4 | 27.0±6.5 | 28.2±6.8 | 26.9±6.5 | <2.20x10<sup>-16</sup> | 26.6±6.7 | 26.7±7.2 | 26.5±6.4 | <2.20x10<sup>-16</sup> |
| **Healthcare workers (%)** | 31,915 (9.5%) | 7,494 (27.6%) | 1190 (58.9%) | 6,304 (25.1%) | 2.94x10<sup>-234</sup> | 5,344 (9.8%) | 1,541 (8.9%) | 3,803 (10.2%) | 1.18x10<sup>-6</sup> |
| **Body rash (%)** | 4,812 (1.4%) | 1,177 (4.3%) | 138 (6.8%) | 1,039 (4.1%) | 1.06x10<sup>-7</sup> | 2,729 (5.0%) | 1,128 (6.5%) | 1,601 (4.3%) | 1.94x10<sup>-20</sup> |
| **Acral rash (%)** | 2,188 (0.6%) | 520 (1.9%) | 62 (3.1%) | 458 (1.8%) | 5.91x10<sup>-5</sup> | 1,210 (2.2%) | 419 (2.4%) | 791 (2.1%) | 0.21 |

Skin-related symptoms were reported by 6,403 users, including 1,534 tested users, and by 3,672 untested symptomatic users. Among the 2,021 users who tested positive on swab test, 178 users (8.8%) reported skin related changes. Of those, 138 (6.8%) reported body rashes and 62 (3.1%) acral rashes (**Table 1)**. Only 22 (1.1%) of them reported both acral and body rashes. Infected users reporting acral rashes were slightly older (mean age = 50.2) than those who did not (mean age = 43.7; Wilcoxon’s test P=6.3×10^−3^). Additionally, prevalence of body rashes was slightly higher among females (OR=1.60, 95% CI=1.08-2.44, P=0.02). We did not observe any significant age difference for body rashes or sex difference for acral rash prevalence (P>0.05).

Similar skin symptoms were also seen in untested symptomatic users: 1,429 (8.2%) users reporting any of the three classic symptoms also reported a rash, compared to 6.0% for those who did not (OR=1.32, 95% CI=1.23-1.42, P<4.7×10^−15^).

Association analysis highlighted higher prevalence of either body or acral rashes among users who tested positive for SARS-CoV-2 compared to those who tested negative (OR=1.67, 95% CI=1.42-1.97, P=1.1×10^− 9^). The subtypes were similar: body rashes were associated with SARS-CoV-2 positive swab with an OR of 1.66 (95% CI=1.37-1.99, P=1.1×10^−9^), whereas the OR for acral rashes was 1.74 (95% CI=1.33-2.28, P=5.9×10^−5^). In comparison, the odd ratio for fever was 1.48 (95% CI=1.31-1.66, P=4.3×10^−11^). Sensitivity analyses confirmed the reported odds ratios (**Supplementary Tables 2** and **3; Supplementary Figures 4** and **5)**. The comparison between 17,371 symptomatic untested users who reported at least one of the classic symptoms and those who did not yielded an OR of 1.46 (95% CI=1.35-1.58, P=1.9×10^−20^) for body rash, while the association with the rarer acral rash was not significant (P=0.21). We could not assess whether ethnicity affected the prevalence of cutaneous symptoms as the number of non-European users with skin symptoms was too low (**Supplementary Tables 1, 2**, and **3, Supplementary Figures 4** and **5**).

To better investigate the duration of skin rashes and their timing in relation to other COVID-19 symptoms, we collected data from 11,544 individuals who responded to an independent on-line survey on possible COVID-19-related skin rashes. Median age was 53 years old (1^st^-3^rd^ quartile: 41-63), 77% were female. Among surveyees, 694 reported a positive SARS-CoV-2 swab or antibody test, and 3,109 were untested but reported having had one of the three classic symptoms. Photos of rashes were shared by 2,328 surveyees, and 260 photos were randomly selected to be assessed by four experienced dermatologists, who divided them in five common categories (*i*.*e*., papular, urticarial, vasculitic body, livedo reticularis, and acral lesions). We discarded 52 photos (20%) rejected by at least one dermatologist because the image was blurred, or the area photographed was too small to evaluate, or the person in the photo could be de-anonymised. Out of the 208 good quality photos, 30 were judged as not attributable to SARS-CoV-2 infection (*e*.*g*., acne, shingles, molluscum contagiosum, pompholyx eczema, peri-oral dermatitis, impetigo, or tinea, 14.4%), and 18 were discarded due to a disagreement in diagnosis between dermatologists (8.7%), resulting in 160 photos used in the subsequent analyses. Notably, when asked whether a photo showed a COVID-19-related skin rash, the four dermatologists agreed 82.2% of the times, and three out of four dermatologists agreed 95.7% of the times. The three most common presentations were papular rashes (including erythemato-papular and erythemato-vesicular types, 41.2%), urticaria (30.0%), and acral lesions (23.1%). Overall agreement for acral lesion, and urticaria were 96.6% and 80.9%, respectively. Papular rashes showed the lowest agreement (76.4%), possibly because some the papular rashes may be more subtle and more difficult to photograph. Examples of COVID-19 cutaneous manifestation are show in **Figure 1 and Supplementary Figures 6-8**, while a large curated library of 400 high quality photos collected *via* the survey is available at https://covidskinsigns.com/.

The average duration of cutaneous rash was 13 days for acral lesions, 14 days for papular, and 5 days for urticaria (significantly shorter duration; Wilcoxon’s P<2.2×10^−6^, **Figure 2**).

The 694 surveyees that reported having tested positive to SARS-CoV-2 *via* a swab or antibody test, with cutaneous signs, also reported other COVID-19-related symptoms, fatigue (11%), headaches (9%), loss of smell (9%), fever (7%), muscle pain (6%), shortness of breath (6%), and persistent cough (6%) being the most common. Interestingly, while most surveyees declared skin changes to appear at the same time as other COVID-19 symptoms (47%) or afterwards (35%), in 17% of the cases skin symptoms appeared before any other symptoms, and in 21% of the cases the rash was the only symptom. Similar estimates were obtained when focusing on the 3,109 symptomatic untested subjects, where 47%, 39%, and 15% surveyees declared to have had cutaneous rash during, after, and before any other symptoms, respectively.

## Discussion

COVID-19 is now known to have varied clinical manifestations and to target multiple organs, including the skin^1–3^. COVID-19 rashes may present in many forms and at different stages of the disease. The heterogeneous presentations and the focus on severely ill patients during the early phases of the pandemic, led to the skin being overlooked as an important target organ for SARS-CoV-2.

In this community-based study, 8.8% of positive COVID-19 cases *via* swab tests also reported skin rashes (OR=1.67, 95% CI=1.42-1.97). Body rashes were more frequent than acral lesions (6.8% *vs* 3.1%) although their predictive value was similar (OR=1.66, 95% CI=1.37-1.99 *vs* 1.74, 95% CI=1.33-2.28, respectively). Interestingly, the odds ratio for both types of rash was greater than for fever (OR=1.48), a widely used criteria to suggest testing. Reports of cases with both body rashes and acral lesions were rare, suggesting different pathogenesis, with the former caused by immunological reactions to the virus, and the latter more likely explained by delayed small thrombotic occlusions or damage to vessel walls^2^.

The use of the app has been valuable to document the presence of different types of COVID-19 symptoms in the community^11,12^. However, data on cutaneous symptoms were only recently collected, and this hindered our ability to identify at which stage of the disease they appear and how long they last. An independent survey was therefore carried out to capture more details on the types of rashes including photos, their duration, timing, results from SARS-CoV-2 swab/antibody test, and co-occurring symptoms. The prevalence of the different types of rashes was assessed by four dermatologists using uploaded photos. This showed that papular rashes were the most frequent, and that urticaria was short lived.

The survey also showed that in 17% of the positive surveyees skin rash was the first symptom to appear, as already observed in two case reports^8,13^, and importantly in 21% of them it was the only symptom, and a SARS-CoV-2 diagnosis would have been missed if using either the UK NHS or the US CDC criteria alone.

### Limitations

A major limitation of this study is the self-reported nature of the data. However, we believe that the presence of a rash, especially if symptomatic, is less subjective and more specific than other symptoms such as fatigue, headaches, or persistent cough. Of the photos uploaded by the surveyees and blindly assessed by four dermatologists, only 14% were classified as definitively non-COVID-19-related dermatological conditions, suggesting that the large majority of surveyees (86%) were able to self-identify cutaneous manifestation likely to be related to COVID-19 infection. This may mean that the total number of COVID-19-related skin rashes self-reported through the app and the survey may have been slightly overestimated. On the other hand, many of the app users may have failed to realise the relevance of cutaneous symptoms and not have reported them if not accompanied by other more known COVID-19 symptoms. Given the very large number of users of both the app and the survey, we are confident that potential errors deriving by the analysis of data reported by 336,847 users are likely reflected by larger standard errors of the estimates, rather than on the point estimate of their effects, as already postulated in other large and successful biobank resources (*e*.*g*., the UK Biobank), which are also based on self-reported health-related data. Second, our study sample is not fully representative of the general population, as it represents a self-selected group of individuals, and also because of the uneven access to SARS-CoV-2 testing in the early stages of the pandemic, with tested subjects encompassing a mixture of healthcare workers, at-risk subjects with chronic diseases, elderly people, etc. Our study sample is also composed predominantly by white individuals, and with a larger proportion of females and younger individuals compared to what was observed in hospital settings. Third, although COVID-19 rashes can be divided into three main types, *i*.*e*., urticarial, erythemato-papular/vesicular, and chilblains/perniosis on acral sites, the app only classified them into two categories, with the urticarial rash and erythemato-papular/vesicular rashes together, as both tend to be itchy and the users may not be able to differentiate between them. Moreover, we did not consider rare dermatological presentations, because the main aim of this study was not to provide an exhaustive description of SARS-CoV-2 cutaneous manifestations but to raise awareness of the high prevalence of common COVID-19 rashes, which can sometimes appear earlier than other COVID-19 symptoms or be the only symptom. To this purpose, we have further developed a large library of curated photos to help GPs and healthcare professionals not specialised in dermatology to recognise the most common categories of COVID-19 rash.

## Conclusions

The NHS in UK lists three main symptoms suspicious of COVID-19 (www.nhs.uk/conditions/coronavirus-covid-19/symptoms), whilst the CDC in the USA lists eleven (https://www.cdc.gov/coronavirus/2019-ncov/symptoms-testing/symptoms.html). However, these do not currently include any skin-related symptoms, although they can be easily spotted by patients. Importantly from the survey reported in this study, it was the only presentation in one fifth of the patients diagnosed with COVID-19.

This study strongly supports the inclusion of skin rashes in the list of suspicious COVID-19 symptoms. Although, it is less prevalent than fever, it is more specific of COVID-19 and lasts longer. An increased awareness from the public and healthcare professionals regarding COVID-19 skin changes will allow more efficient dectection of infection and contact tracing. It is important to increase awareness of the rashes associated with COVID-19 infection. We have established a large library of high quality manually curated photos, available at: https://covidskinsigns.com, to help healthcare professionals to identify COVID-19-related dermatological conditions.

## Data Availability

Data collected in the app are being shared with other health researchers through the NHS-funded Health Data Research UK (HDRUK)/SAIL consortium, housed in the UK Secure e-Research Platform (UKSeRP) in Swansea. Anonymized data collected by the symptom tracker app can be shared with researchers who provide a methodologically sound proposal via HDRUK, provided the request is made according to their protocols and is in the public interest (see https://healthdatagateway.org/detail/9b604483-9cdc-41b2-b82c-14ee3dd705f6). Data updates can be found at https://covid.joinzoe.com. The app code is publicly available from https://github.com/zoe/covid-tracker-react-native. The data screening script is publicly available from https://github.com/KCL-BMEIS/zoe-data-prep.

## Contributors

VB, AV, TDS, and MF conceived the study. JW, CJS, and TDS conceived COVID Symptom Study app. VB, AB, TDS conceived the survey. AV and NR curated the COVID Symptom Study app data with contribution from RCEB, BM and SO. AV, NR, and MF curated the survey data. VB, JK, RM, and SP assessed and categorised the photos. VB, JK, RN curated the British Association of Dermatologists COVID-19 Skin Gallery database. AV, NR, and MF performed the statistical analyses. VB, AV, and MF drafted the manuscript. AV, NR, and MF had access to the raw app and survey data, but not to the photographs. AB, VB, JK, RM, RN, and SP had access to the photographs. All authors had access to the summary statistics. All authors read and approved the final version of the manuscript.

## Data sharing

Data collected in the app are being shared with other health researchers through the NHS-funded Health Data Research UK (HDRUK)/SAIL consortium, housed in the UK Secure e-Research Platform (UKSeRP) in Swansea. Anonymized data collected by the symptom tracker app can be shared with researchers who provide a methodologically sound proposal *via* HDRUK, provided the request is made according to their protocols and is in the public interest (see https://healthdatagateway.org/detail/9b604483-9cdc-41b2-b82c-14ee3dd705f6). Data updates can be found at https://covid.joinzoe.com. The app code is publicly available from https://github.com/zoe/covid-tracker-react-native. The data screening script is publicly available from https://github.com/KCL-BMEIS/zoe-data-prep.

## Acknowledgements

Zoe Global Limited provided in kind support for all aspects of building, running, and supporting the app and service to all users worldwide. Investigators received support from the Wellcome Trust, the MRC/BHF, Alzheimer’s Society, EU, NIHR, CDRF, and the NIHR-funded BioResource, Clinical Research Facility and BRC based at GSTT NHS Foundation Trust in partnership with King’s College London. This work was also supported by the UK Research and Innovation London Medical Imaging & Artificial Intelligence Centre for Value Based Healthcare.

