## Supplementary Figures and Tables for "Diagnostic value of cutaneous manifestation of SARS-CoV-2 infection"

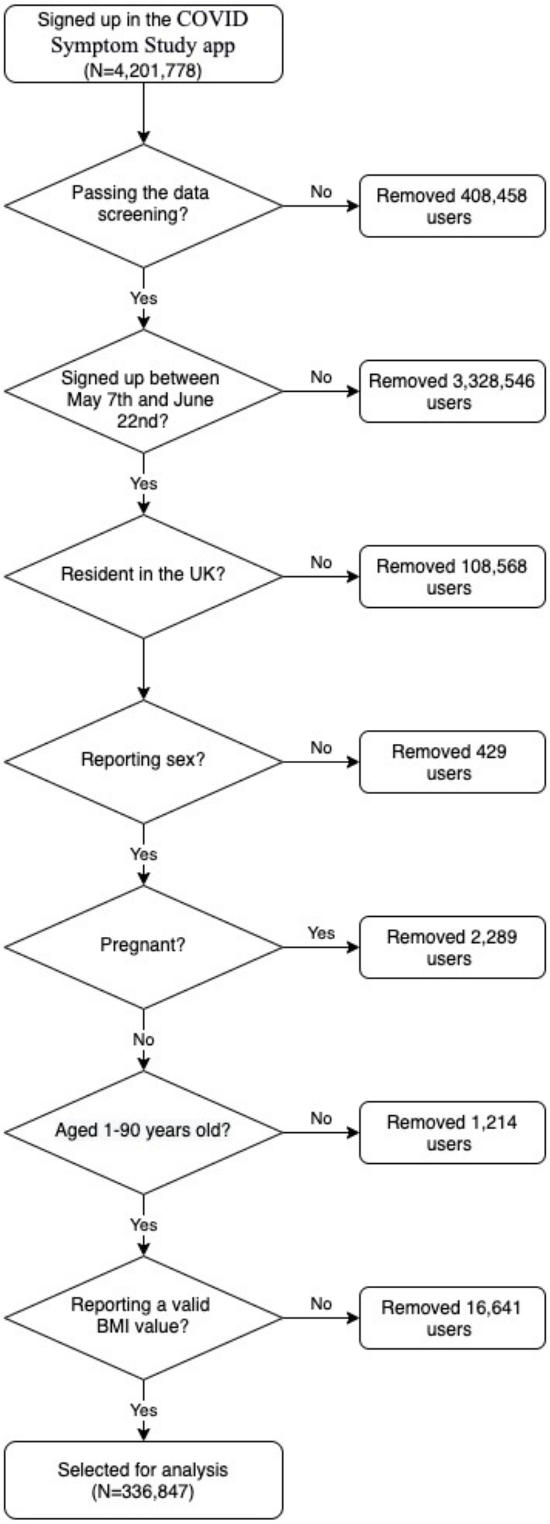


**Supplementary Figure 1.** The flowchart depicts the protocol used for selecting the study sample from the COVID Symptom Study app data. The data screening was carried out using the zoe-data-prep python script (https://github.com/KCL-BMEIS/zoe-data-prep, version 0.1.9) which was used to retain users satisfying the following criteria: logged at least once, age range between 0 and 120 years old, height between 20 and 220 cm, weight between 3 and 200 kg, and body mass index (BMI) between 0 and 100 kg/m$^2$. Valid body mass index (BMI) values were: inside the range of 15 to 55 kg/m$^2$, for users 16 years old or older, or inside two standard deviation from the sample's mean for each age, for users younger than 16 years old.

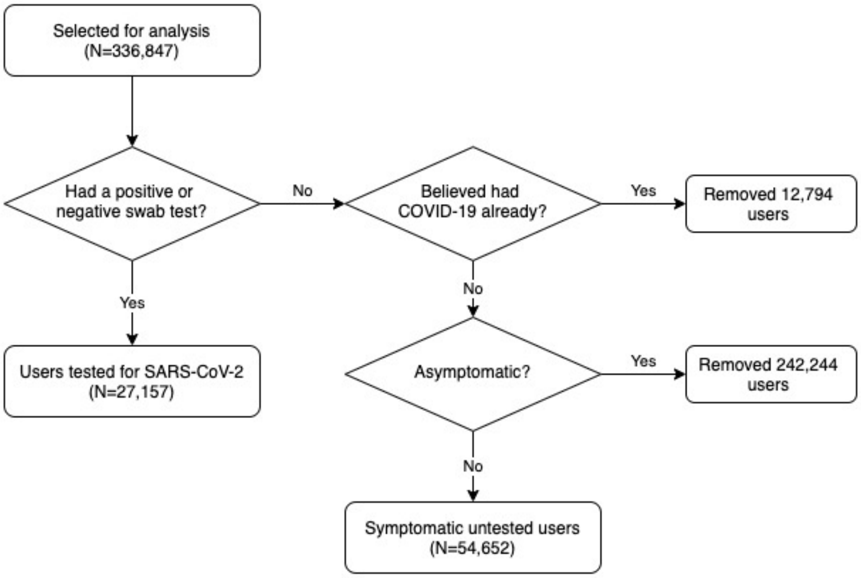


**Supplementary Figure 2.** The flowchart depicts the protocol used to partition the selected COVID Symptom Study app users (see Supplementary Figure 1) in users tested for SARS-CoV-2 and in untested asymptomatic users.

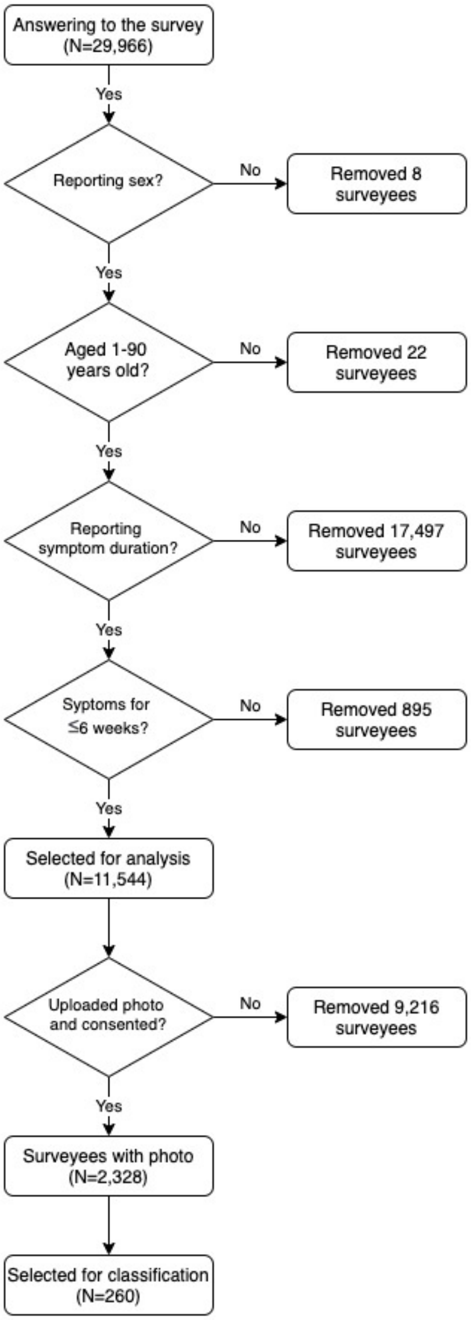


**Supplementary Figure 3.** The flowchart depicts the protocol used for selecting the study sample from the survey data.

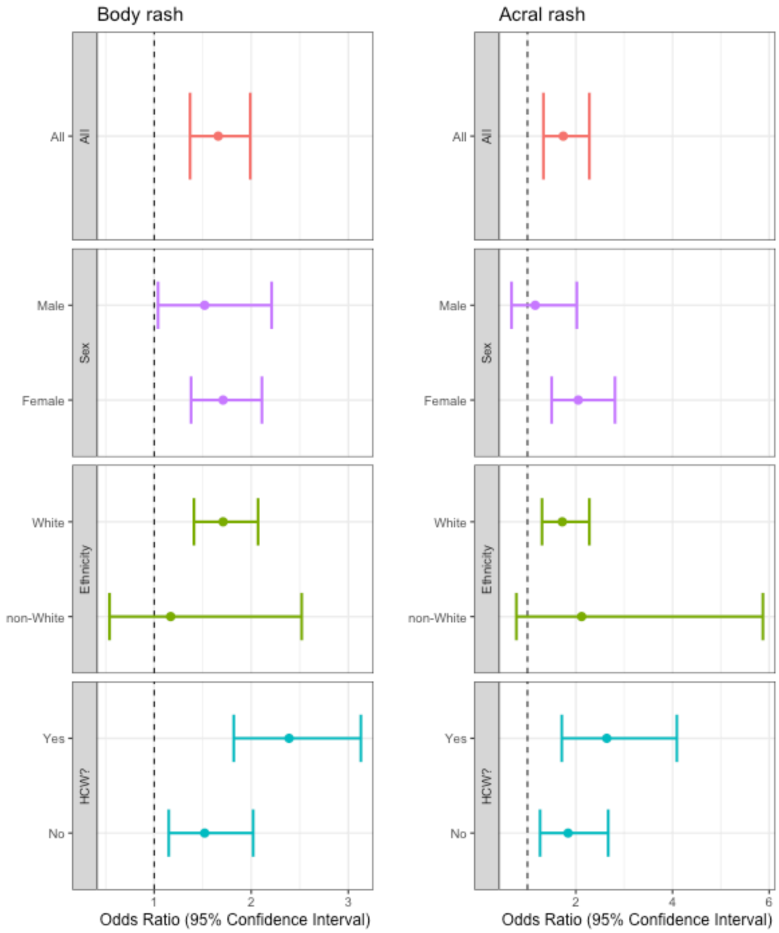


**Supplementary Figure 4.** Sensitivity analysis for multivariate logistic regression in users tested for SARS-CoV-2 infection. Forest plots showing the OR of self-reporting a skin-related symptom in users tested for SARS-CoV-2 infection, stratified by sex, ethnicity, and being a healthcare worker (HCW).

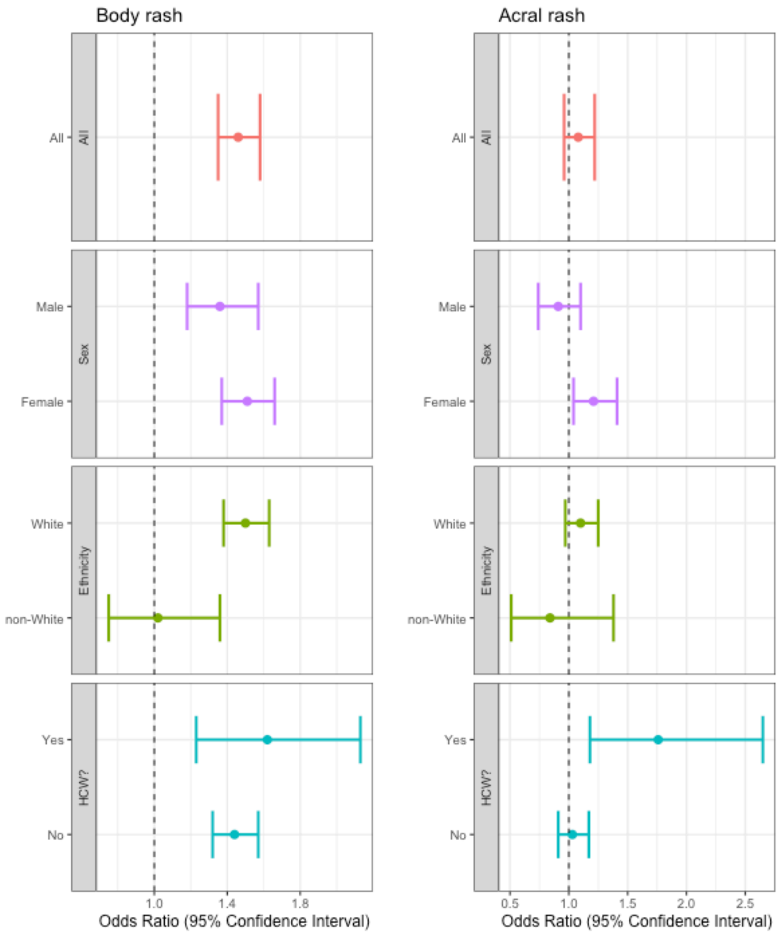


**Supplementary Figure 5.** Sensitivity analysis for multivariate logistic regression in untested symptomatic users. Forest plots showing the OR of self-reporting a skin-related symptom in users while experiencing at least one of the three classic symptoms included in the UK NHS guidelines (*i.e.,* fever, persistent cough, and/or anosmia) stratified by sex, ethnicity, and being a healthcare worker (HCW).

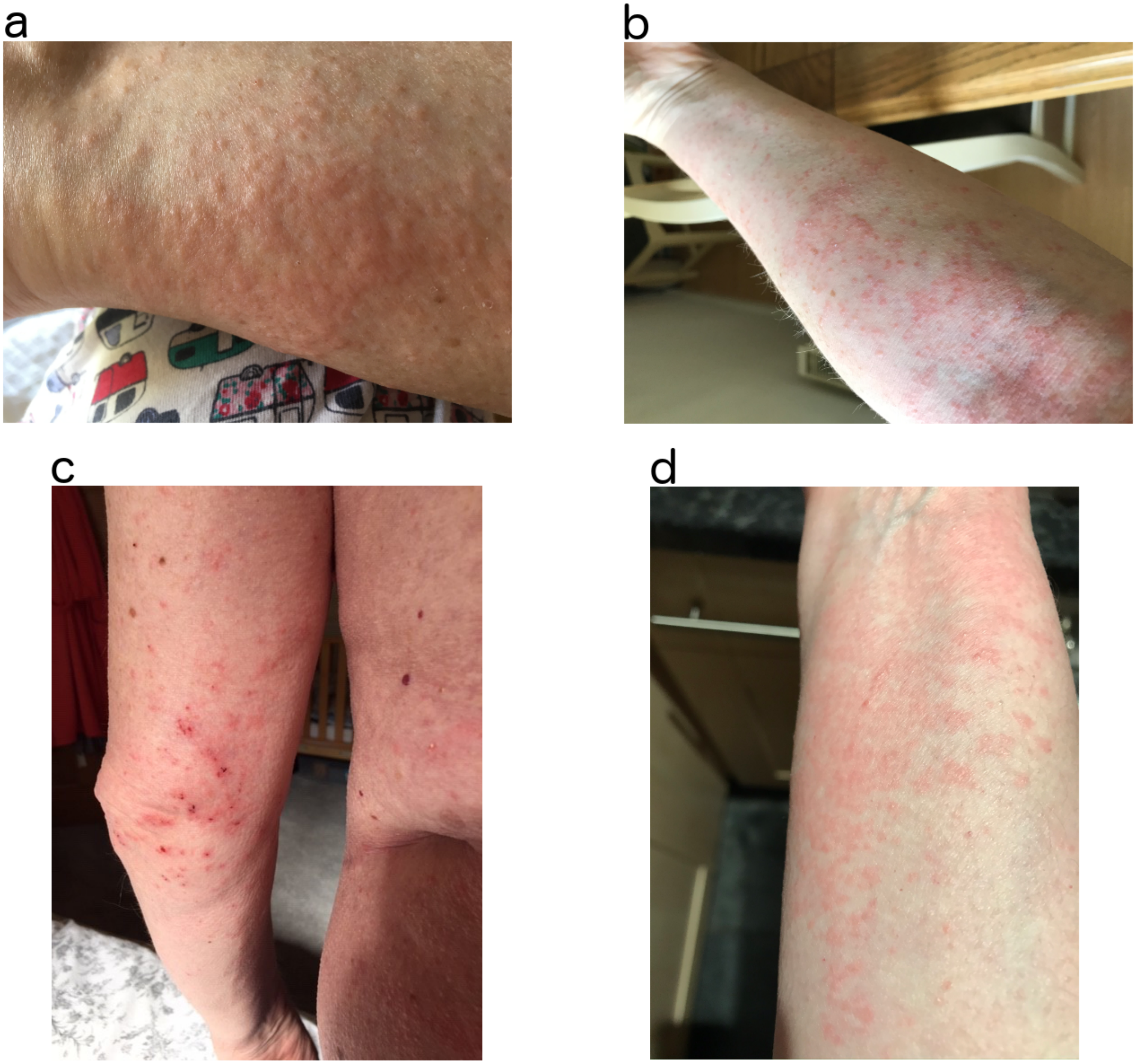

**Supplementary Figure 6.** Examples of papular rash. (a) Erythematopapular rash on forearm. (b) Erythematopapular rash on arm. (c) Erythematopapular rash with some dried vesicles on the elbows. (d) Erythematopapular rash on the forearm.

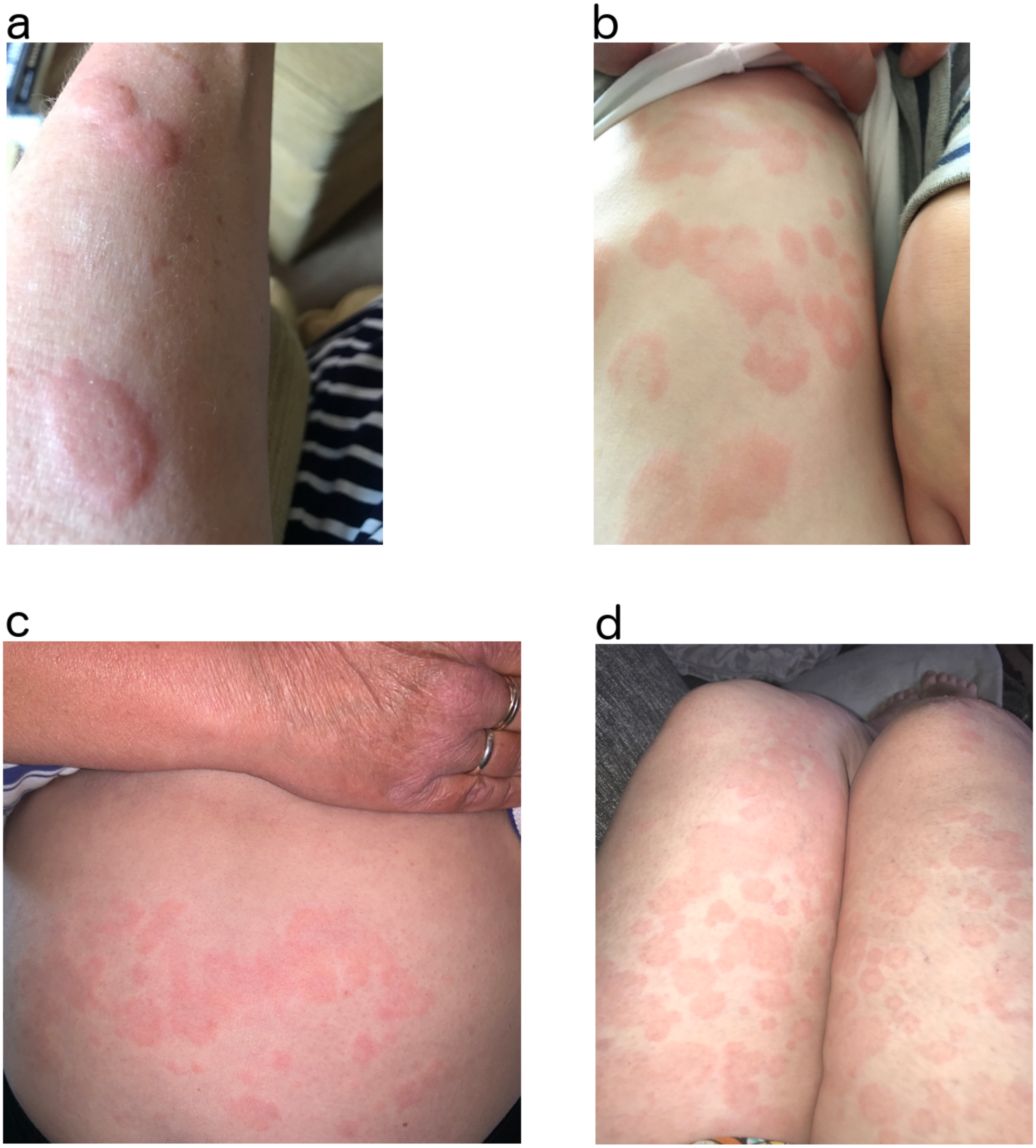


**Supplementary Figure 7.** Examples of urticarial rash. (a) Tumid urticated plaques on arm. (b) Urticated plaques on the abdomen with annular lesions. (c) Urticarial rash on abdomen. (d) Widespread urticaria on the thighs.

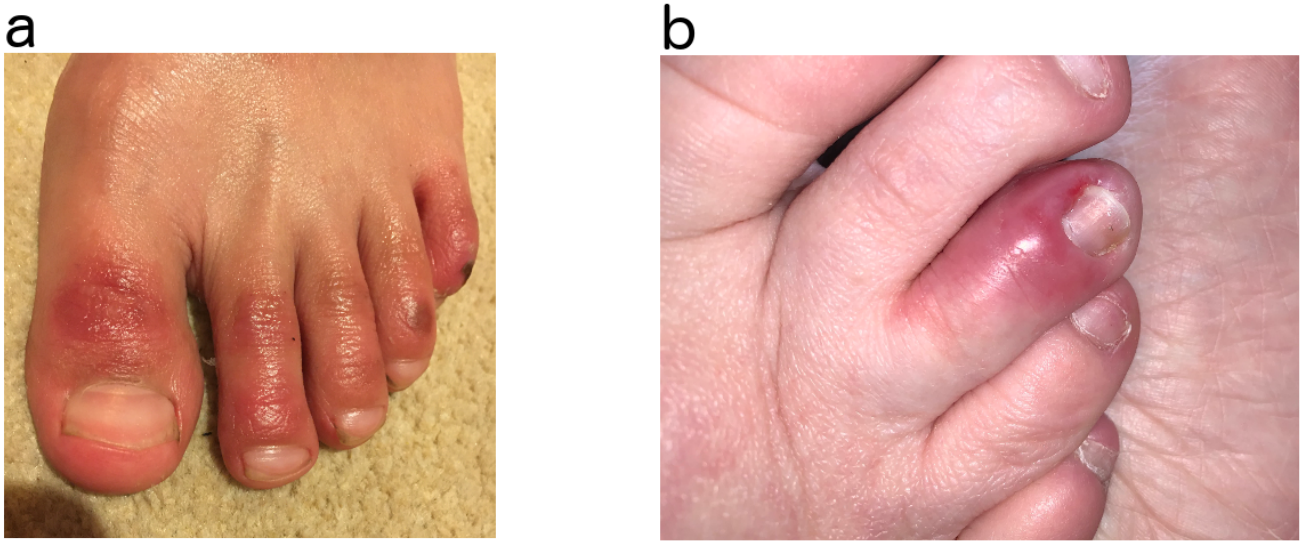


**Supplementary Figure 8.** Examples of acral rash. (a) Erythema on the dordal aspects of all toes with some epidermal necrosis on the 4th and 5th toe. (b) Swelling of the toe with erythema.

### Supplementary Tables

**Supplementary Table 1.** Sample characteristics. Values are reported as number and percentage. Ethnicity, smoking, disease status and medications were compared using Pearson's $\chi^2$ test, as implemented in the prop.test function (stats R package, v 3.6.0). "Users tested for SARS-CoV-2" refers to users self-reporting a positive or negative swab test result. "Symptomatic untested users" refers to users who reported at least one of the 16 collected symptoms, did not believe that they had already had COVID-19 when registering with the app, and had not been tested for SARS-CoV-2. "Classic symptoms" refer to those included in the NHS guidelines (*i.e.,* fever, persistent cough, and/or anosmia), whose presence would require isolation and testing for SARS-CoV-2 infection.

| | | All users | Users tested for SARS-CoV-2 | | | | Symptomatic untested users | | | |
|---|---|---|---|---|---|---|---|---|---|---|
| | | | All | Positive | Negative | P | All | With classical symptoms | Without classic symptoms | P |
| N | | 336,847 | 27,157 | 2,021 | 25,136 | | 54,652 | 17,371 | 37,281 | |
| Ethnicity | | | | | | $4.72 \times 10^{-6}$ | | | | $<2 \times 10^{-16}$ |
| | Asian | 6,503 (1.9%) | 646 (2.4%) | 70 (3.5%) | 576 (2.3%) | | 1,029 (1.9%) | 422 (2.4%) | 607 (1.6%) | |
| | Black | 2,105 (0.6%) | 232 (0.9%) | 34 (1.7%) | 198 (0.8%) | | 370 (0.7%) | 143 (0.8%) | 227 (0.6%) | |
| | Chinese | 1,029 (0.3%) | 87 (0.3%) | 6 (0.3%) | 81 (0.3%) | | 134 (0.2%) | 43 (0.2%) | 91 (0.2%) | |
| | Middle East | 1,204 (0.4%) | 114 (0.4%) | 13 (0.6%) | 101 (0.4%) | | 187 (0.3%) | 84 (0.5%) | 103 (0.3%) | |
| | Mixed | 6,974 (2.1%) | 526 (1.9%) | 35 (1.7%) | 491 (2.0%) | | 1,295 (2.4%) | 508 (2.9%) | 787 (2.1%) | |
| | White | 316,567 (94.0%) | 25,341 (93.3%) | 1,840 (91.0%) | 23,5012 (93.5%) | | 51,188 (93.7%) | 16,021 (92.2%) | 35,167 (94.3%) | |
| | N/A | 2,465 (0.7%) | 211 (0.8%) | 23 (1.1%) | 188 (0.7%) | | 449 (0.8%) | 150 (0.9%) | 299 (0.8%) | |
| Smoking status | | | | | | 0.04 | | | | $<2 \times 10^{-16}$ |
| | Never | 241,093 (71.6%) | 18,499 (68.1%) | 1,396 (69.1%) | 17,103 (68.0%) | | 37,932 (69.4%) | 11,752 (67.7%) | 26,180 (70.2%) | |
| | Ex | 62,563 (18.6%) | 5,655 (20.8%) | 436 (21.6%) | 5,219 (20.8%) | | 10,726 (19.6%) | 3,193 (18.4%) | 7,533 (20.2%) | |
| | Current | 33,176 (9.8%) | 3,002 (11.1%) | 189 (9.4%) | 2,813 (11.2%) | | 5,994 (11.0%) | 2,426 (14.0%) | 3,568 (9.6%) | |

| | | | | | | | | | |
|---|---|---|---|---|---|---|---|---|---|
| Has diabetes (%) | 12,884 (3.8%) | 1,1191 (4.4%) | 113 (5.6%) | 1,078 (4.3%) | $7.04 \times 10^{-3}$ | 2,109 (3.9%) | 704 (4.1%) | 1,405 (3.8%) | 0.11 |
| Has heart disease (%) | 10,052 (3.0%) | 936 (3.4%) | 78 (3.9%) | 858 (3.4%) | 0.32 | 1,663 (3.0%) | 529 (3.0%) | 1,134 (3.0%) | 1.00 |
| Has lung disease (%) | 31,489 (9.3%) | 3,150 (11.6%) | 264 (13.1%) | 2,886 (11.5%) | $3.58 \times 10^{-2}$ | 6,912 (12.6%) | 2,603 (15.0%) | 4,309 (11.6%) | $3.71 \times 10^{-29}$ |
| Has kidney disease (%) | 2,598 (0.8%) | 293 (1.1%) | 29 (1.4%) | 264 (1.1%) | 0.13 | 527 (1.0%) | 169 (1.0%) | 358 (1.0%) | 0.93 |
| Has cancer (%) | 4,456 (1.3%) | 543 (2.0%) | 34 (1.7%) | 509 (2.0%) | 0.33 | 660 (1.2%) | 210 (1.2%) | 450 (1.2%) | 1.00 |
| Corticosteroids (%) | 24,583 (7.3%) | 2,491 (9.2%) | 190 (9.4%) | 2,301 (9.2%) | 0.74 | 5,601 (10.2%) | 2,028 (11.7%) | 3,573 (9.6%) | $6.95 \times 10^{-14}$ |
| Immunosuppressants (%) | 12,006 (3.6%) | 1,138 (4.2%) | 75 (3.7%) | 1,063 (4.2%) | 0.29 | 2,415 (4.4%) | 910 (5.2%) | 1,505 (4.0%) | $2.26 \times 10^{-10}$ |
| Blood pressure medications (%) | 44,061 (13%) | 3,593 (13.2%) | 273 (13.5%) | 3,320 (13.2%) | 0.73 | 6,272 (11.5%) | 1.852 (10.7%) | 4,420 (11.9%) | $4.80 \times 10^{-5}$ |

**Supplementary Table 2.** Sensitivity analysis for multivariate logistic regression in users tested for SARS-CoV-2 infection. Associations between the presence/absence of self-reported skin-related symptoms and test results in users tested for SARS-CoV-2 infection, stratified by sex, ethnicity and being a healthcare worker, were carried out through multivariate logistic regression, and the following variables were included as covariates: sex, age, BMI, ethnicity (namely: Asian, Black, Chinese, Middle Eastern, White, or mixed), smoking status (namely: never, ex, current), common disease status (namely: diabetes and lung disease), and whether corticosteroids, immunosuppressants, or blood pressure medications were administered.

| | | | **Body rash** | | | **Acral Rash** | | |
|---|---|---|---|---|---|---|---|---|
| **Risk factor** | **Stratum** | **N** | **OR** | **95% CI** | **P** | **OR** | **95% CI** | **P** |
| | All sample | 26,945 | 1.66 | 1.37-1.99 | $1.06 \times 10^{-7}$ | 1.74 | 1.33-2.28 | $5.91 \times 10^{-5}$ |
| Sex | Male | 10,598 | 1.52 | 1.04-2.21 | 0.03 | 1.16 | 0.67-2.02 | 0.590 |
| | Female | 16,347 | 1.71 | 1.38-2.11 | $1.11 \times 10^{-6}$ | 2.05 | 1.50-2.81 | $7.69 \times 10^{-6}$ |
| Ethnicity | White | 25,340 | 1.71 | 1.41-2.07 | $4.51 \times 10^{-8}$ | 1.72 | 1.30-2.28 | $1.71 \times 10^{-4}$ |
| | Non-White | 1,605 | 1.17 | 0.54-2.52 | 0.698 | 2.12 | 0.77-5.87 | 0.146 |
| Healthcare worker? | Yes | 7,429 | 2.39 | 1.82-3.13 | $2.87 \times 10^{-10}$ | 2.64 | 1.71-4.09 | $1.27 \times 10^{-5}$ |
| | No | 19,516 | 1.52 | 1.15-2.02 | $3.44 \times 10^{-3}$ | 1.84 | 1.26-2.67 | $1.45 \times 10^{-3}$ |

**Supplementary Table 3.** Sensitivity analysis for multivariate logistic regression in untested symptomatic users. Associations between the presence/absence of self-reported skin-related symptoms and presence/absence of at least one the three classic symptoms included in the NHS guidelines (*i.e.,* fever, persistent cough, and/or anosmia) in untested symptomatic users, stratified by sex, ethnicity and being a healthcare worker, were carried out through multivariate logistic regression, and the following variables were included as covariates: sex, age, BMI, ethnicity (namely: Asian, Black, Chinese, Middle Eastern, White, or mixed), smoking status (namely: never, ex, current), common disease status (namely: diabetes and lung disease), and whether corticosteroids, immunosuppressants, or blood pressure medications were administered.

| | | | **Body rash** | | | **Acral Rash** | | |
|---|---|---|---|---|---|---|---|---|
| **Risk factor** | **Stratum** | **N** | **OR** | **95% CI** | **P** | **OR** | **95% CI** | **P** |
| | All sample | 54,203 | 1.46 | 1.35-1.58 | $1.94 \times 10^{-20}$ | 1.08 | 0.96 1.22 | 0.205 |
| Sex | Male | 19,697 | 1.36 | 1.18-1.57 | $1.46 \times 10^{-5}$ | 0.91 | 0.74 1.10 | 0.331 |
| | Female | 34,506 | 1.51 | 1.37-1.66 | $2.34 \times 10^{-16}$ | 1.21 | 1.04 1.41 | 0.016 |
| Ethnicity | White | 51,188 | 1.50 | 1.38-1.63 | $6.36 \times 10^{-22}$ | 1.10 | 0.97 1.25 | 0.135 |
| | Non-White | 3,015 | 1.02 | 0.75-1.36 | 0.921 | 0.84 | 0.51 1.38 | 0.486 |
| Healthcare worker? | Yes | 5,298 | 1.62 | 1.23-2.13 | $5.34 \times 10^{-4}$ | 1.76 | 1.18 2.65 | $6.12 \times 10^{-3}$ |
| | No | 48,905 | 1.44 | 1.32-1.57 | $1.47 \times 10^{-17}$ | 1.03 | 0.91 1.17 | 0.631 |
